# Exploring Sleep Quality in Bangladeshi University Students during the COVID-19 Pandemic

**DOI:** 10.1101/2025.04.06.25325308

**Authors:** M Nurunnabi, MK Khan, FR Kaiser, MG Abbas, TT Tabassum, T Farha, F Akter, MA Tarafdar

## Abstract

**Background:** University students frequently experience challenges that affect their sleep quality, including academic stress, lifestyle habits, and socio-demographic factors. Common sleep disturbances, such as delayed sleep onset and poor sleep efficiency, often result in daytime dysfunction and difficulty maintaining enthusiasm.

**Methods:** This cross-sectional study appraised sleep quality among undergraduate students in Bangladesh during the COVID-19 pandemic. A total of 530 undergraduate students were purposively selected from three public universities. Participants were chosen through convenience sampling, with students who had contracted COVID-19 during the study excluded from the sample.

**Results:** The mean age of the university students was 21.1 (±2.0) years. Regarding PSQI components, the participants showed moderate sleep latency (55.7%) and habitual sleep efficiency (56.0%), with mild levels of sleep disturbances at night (68.1%), daytime dysfunction (51.1%), and difficulty maintaining enthusiasm (48.5%). The global PSQI scores were poor for 59.4% of participants, while 40.6% had good scores, with a mean of 6.5(±2.9). Among the students, 44.9% reported fair subjective sleep quality, followed by good (24.7%), very good (18.1%), and poor (12.3%) sleep quality. Lifestyle factors such as class attendance, online media usage, BMI, physical exercise, tea/coffee consumption, and smoking were significantly associated with sleep quality (p<0.05). Sleep quality factors like daytime sleep duration, nighttime sleep latency, and sleeping pill usage were also significantly associated with sleep quality (p<0.05). Significant associations were found between sleep quality and factors such as age, household income, daily study hours, BMI, daytime sleep, and sleep latency (p<0.05).

**Conclusion:** The study revealed that longer sleep latency, daytime sleep, and sleeping pill usage were related with poorer sleep quality. It also emphasized that sleep disturbances, daytime dysfunction, and difficulty maintaining enthusiasm were significant predictors of sleep quality.

## Introduction

Sleep is an essential physiological function for general health and wellbeing^1^. It is crucial for preserving the best possible health and performance since it is important for many facets of physical, mental, and emotional functioning^2^. Numerous advantages of getting enough sleep include enhanced mood control, cognitive function, immune system performance, and metabolic health^3^. Good sleep habits are essential for self-regulation, the brain’s executive function that governs our actions and a deficit of these habits can result from persistently interrupting the body’s natural sleep cycle or from failing to initiate sleep, which wears one out physically, mentally, and emotionally^4^.

Sleep quality sorting, particularly using indices comprised of the Pittsburgh Sleep Quality Index (PSQI), offers a comprehensive assessment of all aspects of sleep, including both subjective and objective information. The PSQI assesses seven categories of sleep quality over a period of one month: subjective sleep quality, sleep latency, sleep duration, habitual sleep efficiency, sleep disturbances, use of sleep medication, and daytime dysfunction^5^. The PSQI is a standardized and accurate measure of sleep quality across groups that are widely used in clinical and research contexts^6^. Furthermore, the PSQI helps identify individual sleep disruptions and their impact on total sleep quality, allowing for focused interventions to enhance sleep outcomes^7^. A study conducted among undergraduate students found that 70.6% reported sleeping less than eight hours, with an average sleep duration of 7.02 hours, indicating that most students experience sleep deprivation^1^. A significant number of college students experience sleep problems, such as poor sleep quality, symptoms of insomnia, and inconsistent sleep patterns^8,9^.

Sleep deprivation has been associated with increased susceptibility to infections and lower vaccine efficacy, underscoring the need of adequate sleep for immunological protection^11^. It has also been demonstrated to impair attention, concentration, decision-making, and problem-solving abilities, hence affecting academic and vocational performance, as sleep have a strong link to cognitive functioning^7,12^. Furthermore, sleep affects mood regulation and emotional well-being. Adequate sleep is necessary for emotional regulation, stress management, and psychological resilience^13^. Sleep disruptions are intimately associated with mood disorders such as depression and anxiety, and sleep therapies are frequently included in treatment plans^14^. It has also an important role in metabolic control and weight management. Insufficient sleep disturbs hormonal balance, causing changes in appetite-regulating hormones like leptin and ghrelin, which can contribute to weight gain and obesity^15,16^. Depression and anxiety symptoms are prevalent among university students who experience poor sleep, and these symptoms are often linked to lower life satisfaction levels^17-19^. Additionally, there is also evidence suggesting that individuals who experience significant psychological distress during their time as students are likely to continue experiencing high levels of distress in their life^20-22^.

Engaging in intentional late-night activities is one facet of this problem^19^. Students’ diverse habits, including excessive use of media devices, smoking, and consumption of caffeine, collectively contribute to poor sleep hygiene^23,24^. Research on sleep quality among Bangladeshi university students is limited, with existing studies primarily exploring links to psychological distress, internet addiction, and the COVID-19 pandemic^25^. This study aims to assess sleep quality among students in two Bangladeshi cities and identify associated factors.

## Methods

### Study design and settings

This cross-sectional study assessed sleep quality among undergraduate students in Bangladesh during the COVID-19 pandemic. Participants were purposively selected from three public universities: Dhaka University, Jahangirnagar University in Dhaka, and Shahjalal University of Science and Technology in Sylhet.

### Sample selection criteria

A total of 530 undergraduate students, aged 18 or older and in their first to fourth year, were selected through convenience sampling. Students who had contracted COVID-19 during the study were excluded. The sample size was calculated using a 95% confidence interval (CI) and a 5% relative precision.

### Data collection methods

Data were collected using a semi-structured questionnaire, pretested with 55 undergraduate students from a private university in Sylhet. Participants completed the questionnaire via Google Forms between July and December 2021, during their enrollment in the study. The questionnaire was designed to include sociodemographic details, lifestyle and sleep quality-related information, and the PSQI scale.

A. **Semi-structured questionnaire for the sociodemographic outlines:** The questionnaire included sociodemographic variables such as age, gender, marital status, education level, residence, monthly income, family history, and COVID-19 infection status.
B. **The Pittsburg Sleep Quality Index (PSQI):** The PSQI in this survey assessed sleep quality across seven areas: sleep duration, disturbance, latency, daytime dysfunction, efficiency, subjective quality, and medication use. Scores ranged from ‘0’ (no difficulty) to ‘3’ (severe difficulty) per area. A global score ≤5 indicated good sleep quality, while >5 indicated poor sleep quality.

### Statistical analysis

Data were entered and analyzed using IBM SPSS Version 26(New York, USA). Descriptive statistics were presented as frequencies (percentages) for categorical data and means (±SD) for continuous data. The Chi-square (χ2) test and independent sample t-test were used to assess associations. Univariate linear regression and logistic regression analyses were performed to test for significance. A p-value <0.05 at a 95% confidence interval was considered statistically significant.

### Ethical aspects

Participation was voluntary, with confidentiality ensured through individual code numbers. Informed consent was obtained after explaining the study’s objectives and potential outcomes. The research adhered to the 2013 revised Declaration of Helsinki and its amendments, or comparable ethical standards. Ethical approval was granted by Sylhet Women’s Medical College (SWMC), Sylhet, Bangladesh (Reference: SWMC/Eth.C/IRB/202145).

## Results

Table I showed that the mean age of the participants was 21.1 (±2.0) years, and one-third (33.6%) of the participants were from the 22-23 age group, while a significant proportion (30.8%) belonged to the adolescent group aged 18-19 years. The male-to-female ratio was nearly equal (1:1.2), and a substantial portion (8.1%) were married. The majority of the primary earners in the families were employed in service-related occupations (61.9%), while smaller proportions (7.0%) were engaged in other occupations, including agricultural work and day labor. The mean of family size was 5.0 (±1.2) persons, with nuclear families and rural residency comprising more than two-thirds (69.9% and 69.4%, respectively). The monthly household income was estimated 42,469.8 (±23,261.4) Bangladeshi Taka (BDT).

**Table I:**
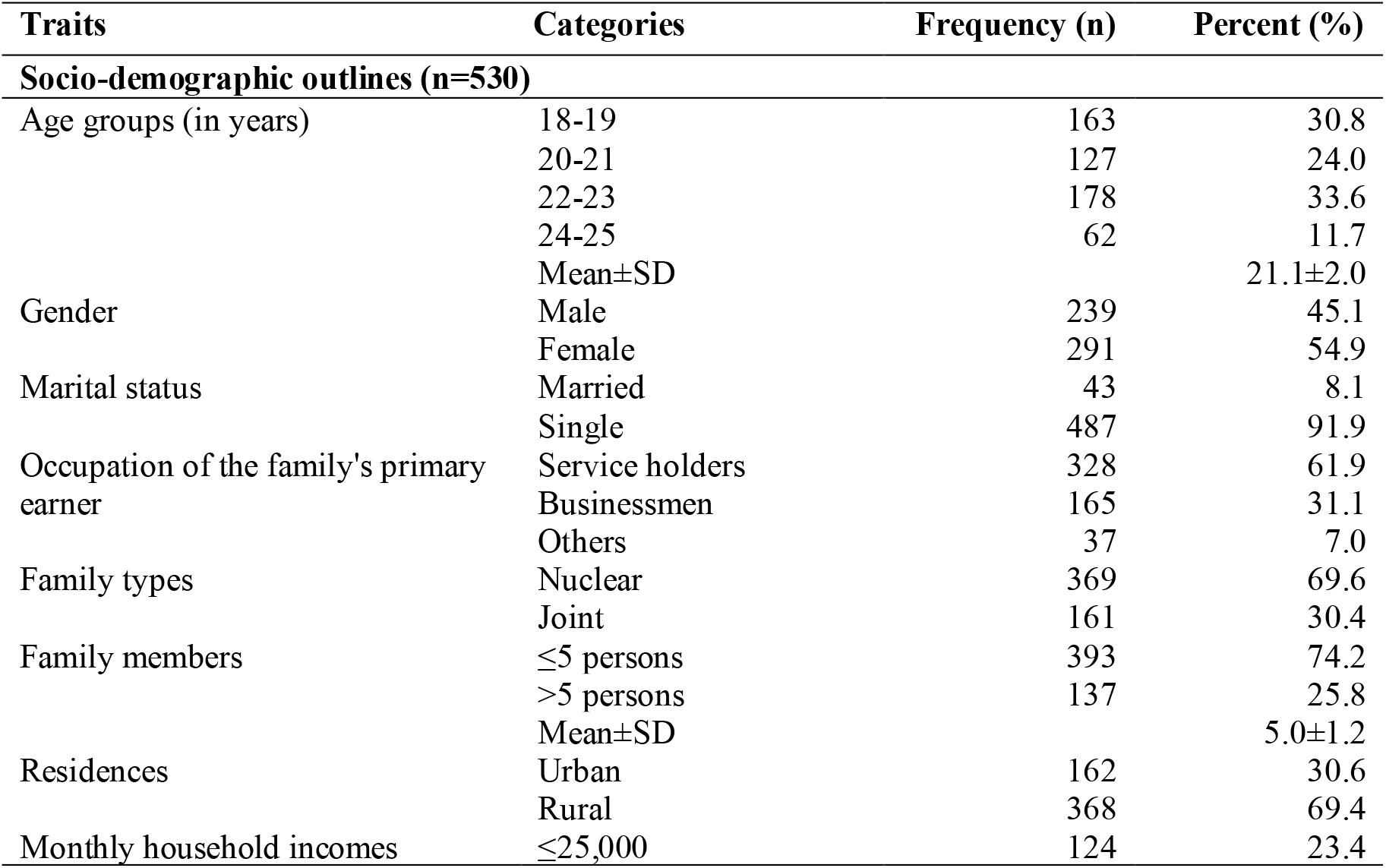
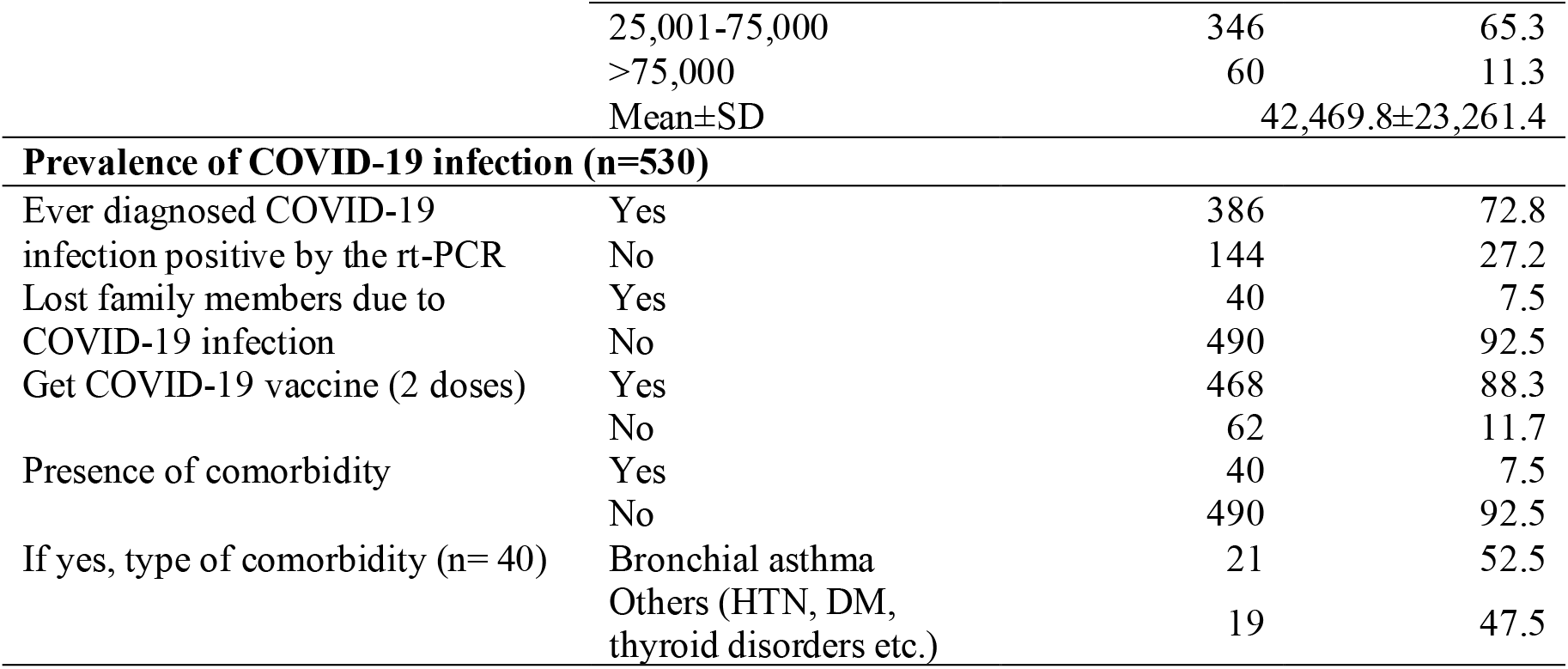
Socio-demographic outlines of students (n=530)

The prevalence of COVID-19 infection among the students was 72.8%, as confirmed by RT-PCR testing. A significant proportion (7.5%) of students reported the loss of second-degree family members due to COVID-19. The majority of students (88.3%) received the second dose of the COVID-19 vaccine, while the remainder did not due to delays in the registration process and the unavailability of the vaccine at health facilities in their areas of residence. Bronchial asthma was the most prevalent medical condition, affecting 52.5% of students with comorbidities, which were present in 7.5% of the students.

Table II described that most students (68.3%) attended online classes regularly, studied ≤4 hours daily (66.8%, mean 3.9±1.9 hours), and spent >4 hours on online media (68.1%, mean 5.4±2.2 hours). The pattern of time spent on online media increased among four-fifths (82.8%) of the students. Nearly one-third of students (31.7%) had a BMI above the normal range (mean 23.5±3.8), while 34.7% engaged in physical exercise regularly, with 54.3% exercising for more than 30 minutes daily. Tea or coffee drinking was frequent among university students (67.9%), with a high prevalence of nighttime consumption (73.9%). Among the students, 40.6% were smokers, with 87.0% of them consuming ≤5 cigarettes per day. The pattern of smoking increased among the majority of respondents, reaching 67.4%.

**Table II:** Factors influences the lifestyle of students during COVID-19 pandemic (n= 530)

| Variables | Categories | Frequency (n) | Percent (%) |
| --- | --- | --- | --- |
| **Lifestyle related factors** |  |  |  |
| Attended online classes | Regularly | 362 | 68.3 |
|  | Irregularly | 168 | 31.7 |
| Studied hours in a day | ≤4 hours | 354 | 66.8 |
|  | >4 hours | 176 | 33.2 |
|  | Mean±SD |  | 3.9±1.9 |
| Spent times on online media | ≤4 hours | 169 | 31.9 |
|  | >4 hours | 361 | 68.1 |
|  | Mean±SD |  | 5.4±2.2 |
| Pattern of spent times on online media | Increased | 439 | 82.8 |
|  | Decreased | 15 | 2.8 |
|  | Didn't change | 76 | 14.3 |
| Body mass index (Kg/m<sup>2</sup>) | <18.5 | 26 | 4.9 |
|  | 18.5- 24.9 | 336 | 63.4 |
|  | 25.0- 29.9 | 96 | 18.1 |
|  | 30.0- 34.9 | 72 | 13.6 |
|  | Mean±SD |  | 23.5±3.8 |
| Performed physical exercises daily | Regularly | 184 | 34.7 |
|  | Occasionally | 346 | 65.3 |
| Duration of exercise (n=184) | ≤30 minutes | 84 | 45.7 |
|  | >30 minutes | 100 | 54.3 |
| Consumption of tea or coffee | Regularly | 360 | 67.9 |
|  | Occasionally | 170 | 32.1 |
| Cups of tea or coffee in a day (n= 360) | ≤2 | 326 | 90.6 |
|  | ≥3 | 34 | 9.4 |
| Consumption of tea or coffee at night (n= 360) | Yes | 266 | 73.9 |
|  | No | 94 | 26.1 |
| History of smoking | Yes | 215 | 40.6 |
|  | No | 315 | 59.4 |
| Number of cigarettes smoked per day (n= 215) | ≤5 sticks | 187 | 87.0 |
|  | >5 sticks | 28 | 13.0 |
| Pattern of smoking (n= 215) | Increased | 145 | 67.4 |
|  | Decreased | 20 | 9.3 |
|  | Didn't change | 50 | 23.3 |
| History of alcohol consumption | Yes | 32 | 6.0 |
|  | No | 498 | 94.0 |
| **Sleep quality related factors** |  |  |  |
| Sleep duration (in a day) | ≤6 hours | 387 | 73.0 |
|  | >6 hours | 143 | 27.0 |
|  | Mean±SD |  | 5.8±1.6 |
| Sleep at daytime | Yes | 358 | 67.5 |
|  | No | 172 | 32.5 |
| Duration of sleep at daytime<br>(n= 358) | ≤2 hours | 143 | 39.9 |
|  | >2 hours | 215 | 60.1 |
|  | Mean±SD |  | 2.7±1.0 |
| Sleep latency at night | ≤30 minutes | 98 | 18.5 |
|  | 31-60 minutes | 85 | 16.0 |
|  | >60 minutes | 347 | 65.5 |
|  | Mean±SD |  | 101.1±47.8 |
| Usage of sleeping pills in the<br>past month | No tablet | 394 | 74.3 |
|  | Occasionally | 94 | 17.7 |
|  | Regularly | 42 | 7.9 |

**Table III:** Association of different factors with the students’ levels sleep quality (n= 530)

| Variables | Categories | N | Levels of sleep quality | | $\chi^2$ value | p-value |
| --- | --- | --- | --- | --- | --- | --- |
|  |  |  | Good | Poor |  |  |
|  |  |  | n(%) | n(%) |  |  |
| Socio-demographic outlines |  |  |  |  |  |  |
| Age groups (years) | 18-19 | 163 | 28(17.2) | 135(82.8) | 69.142 | 0.000 |
|  | 20-21 | 127 | 50(39.4) | 77(60.6) |  |  |
|  | 22-23 | 178 | 109(61.2) | 69(38.8) |  |  |
|  | 24-25 | 62 | 28(45.2) | 34(54.8) |  |  |
| Occupation of the family's primary earner | Service holders | 328 | 70(21.3) | 258(78.7) | 136.163 | 0.000 |
|  | Businessmen | 165 | 124(75.2) | 41(24.8) |  |  |
|  | Others | 37 | 21(56.8) | 16(43.2) |  |  |
| Residences | Urban | 162 | 80(49.4) | 82(50.6) | 7.522 | 0.007 |
|  | Rural | 368 | 135(36.7) | 233(63.3) |  |  |
| Type of family | Nuclear | 369 | 162(43.9) | 207(56.1) | 5.608 | 0.021 |
|  | Joint | 161 | 53(32.9) | 108(67.1) |  |  |
| Monthly household incomes (BDT) | ≤25,000 | 124 | 83(66.9) | 41(33.1) | 57.426 | 0.000 |
|  | 25,001-75,000 | 346 | 124(35.8) | 222(64.2) |  |  |
|  | >75,000 | 60 | 8(13.3) | 52(86.7) |  |  |
| Prevalence of COVID-19 infection |  |  |  |  |  |  |
| Ever diagnosed COVID-19 positive | Yes | 386 | 113(29.3) | 273(70.7) | 75.128 | 0.000 |
|  | No | 144 | 102(70.8) | 42(29.2) |  |  |
| Lost family members due to COVID-19 infection | Yes | 40 | 6(15.0) | 34(85.0) | 11.729 | 0.001 |
|  | No | 490 | 209(42.7) | 281(57.3) |  |  |
| Presence of comorbidity | Yes | 40 | 10(25.0) | 30(75.0) | 4.348 | 0.044 |
|  | No | 490 | 205(41.8) | 285(58.2) |  |  |
| Lifestyle related factors |  |  |  |  |  |  |
| Attended online classes | Regularly | 362 | 186(51.4) | 176(48.6) | 55.405 | 0.000 |
|  | Irregularly | 176 | 29(17.3) | 139(82.7) |  |  |
| Pattern of spent times on online media | Increased | 439 | 194(44.2) | 245(55.8) | 14.645 | 0.001 |
|  | Decreased | 15 | 2(13.3) | 13(86.7) |  |  |
|  | Didn't change | 76 | 19(25.0) | 57(75.0) |  |  |
| Body mass index | <18.5 | 26 | 4(15.4) | 22(84.6) | 76.078 | 0.000 |
|  | 18.5- 24.9 | 336 | 183(54.5) | 153(45.5) |  |  |
|  | 25.0- 29.9 | 96 | 21(21.9) | 75(78.1) |  |  |
|  | 30.0- 34.9 | 72 | 7(9.7) | 65(90.3) |  |  |
| Performed physical exercises | Regularly | 184 | 132(71.7) | 52(28.3) | 113.600 | **0.000** |
|  | Occasionally | 346 | 83(24.0) | 263(76.0) |  |  |
| Consumption of tea or coffee | Regularly | 360 | 122(33.9) | 238(66.1) | 20.755 | **0.000** |
|  | Occasionally | 170 | 93(54.7) | 77(45.3) |  |  |
| Pattern of smoking (n= 215) | Increased | 145 | 54(37.2) | 91(62.8) | 27.471 | **0.000** |
|  | Decreased | 20 | 19(95.0) | 1(5.0) |  |  |
|  | Didn't change | 50 | 15(30.0) | 35(70.0) |  |  |
| **Sleep quality related factors** |  |  |  |  |  |  |
| Duration of sleep at daytime (n= 358) | ≤2 hours | 143 | 70(49.0) | 73(51.0) | 6.579 | **0.012** |
|  | >2 hours | 215 | 76(35.3) | 139(64.7) |  |  |
| Sleep latency at night | ≤30 minutes | 98 | 77(78.6) | 21(21.4) | 77.920 | **0.000** |
|  | 31-60 minutes | 85 | 37(43.5) | 48(56.5) |  |  |
|  | >60 minutes | 347 | 101(29.1) | 246(70.9) |  |  |
| Usage of sleeping pills in the past month | No tablet | 394 | 189(48.0) | 205(52.0) | 52.103 | **0.000** |
|  | Occasionally | 94 | 7(7.4) | 87(92.6) |  |  |
|  | Regularly | 42 | 19(45.2) | 53(54.8) |  |  |
| **The PSQI components** |  |  |  |  |  |  |
| Sleep latency | Normal | 75 | 70(93.3) | 5(6.7) | 112.450 | **0.000** |
|  | Mild | 84 | 31(36.9) | 53(63.1) |  |  |
|  | Moderate | 295 | 103(34.9) | 192(65.1) |  |  |
|  | Severe | 76 | 11(14.5) | 65(85.5) |  |  |
| Habitual sleep efficiency | Normal | 109 | 95(87.2) | 14(12.8) | 150.984 | **0.000** |
|  | Mild | 93 | 12(12.9) | 81(87.1) |  |  |
|  | Moderate | 297 | 108(36.4) | 189(63.6) |  |  |
|  | Severe | 31 | 0(0.0) | 31(100) |  |  |
| Sleeping disturbance at night | Normal | 115 | 47(40.9) | 68(59.1) | 19.658 | **0.000** |
|  | Mild | 361 | 161(44.6) | 200(55.4) |  |  |
|  | Moderate | 36 | 4(11.1) | 32(88.9) |  |  |
|  | Severe | 18 | 3(16.7) | 15(83.3) |  |  |
| Daytime dysfunction | Normal | 151 | 60(39.7) | 91(60.3) | 95.621 | **0.000** |
|  | Mild | 271 | 153(56.5) | 118(43.5) |  |  |
|  | Moderate | 80 | 2(2.5) | 78(97.5) |  |  |
|  | Severe | 28 | 0(0.0) | 28(100) |  |  |
| Problem keeping up enthusiasm daytime | Normal | 153 | 62(40.5) | 91(59.5) | 89.776 | **0.000** |
|  | Mild | 257 | 146(56.8) | 111(43.2) |  |  |
|  | Moderate | 74 | 1(1.4) | 73(98.4) |  |  |
|  | Severe | 46 | 6(13.0) | 40(87.0) |  |  |
| Levels of SSQ | Very good | 96 | 78(81.3) | 18(18.8) | 391.693 | **0.000** |
|  | Good | 131 | 124(94.7) | 7(5.3) |  |  |
|  | Fair | 238 | 6(2.5) | 232(97.5) |  |  |
|  | Poor | 65 | 7(10.8) | 58(89.2) |  |  |

Chi-square test done, $p < 0.05$ considered as statistically significant value

The sleep duration was ≤6 hours daily for three-fourths (73.0%), with a mean of 5.8 (±1.6) hours. Nearly two-thirds of the students (67.5%) slept during the daytime, with a mean duration of 2.7 (±1.0) hours. The mean duration of sleep latency at night was 101.1 (±47.8) minutes. A significant proportion (7.9%) used to tale sleeping pills regularly, and 17.7% used them occasionally.

In terms of PSQI components, it showed that the participants had a moderate level of sleep latency (55.7%) and habitual sleep efficiency (56.0%), with a mild level of sleeping disturbance at night (68.1%), daytime dysfunction (51.1%), and difficulty maintaining enthusiasm during the day (48.5%) (figure 1). The global PSQI scores were poor for 59.4% of participants, while 40.6% had good scores, with a mean of 6.5 (±2.9) (figure 2). Among the students, 44.9% reported fair subjective sleep quality (SSQ), followed by good (24.7%), very good (18.1%), and poor (12.3%) sleep quality (figure 3).

**Figure 1:**
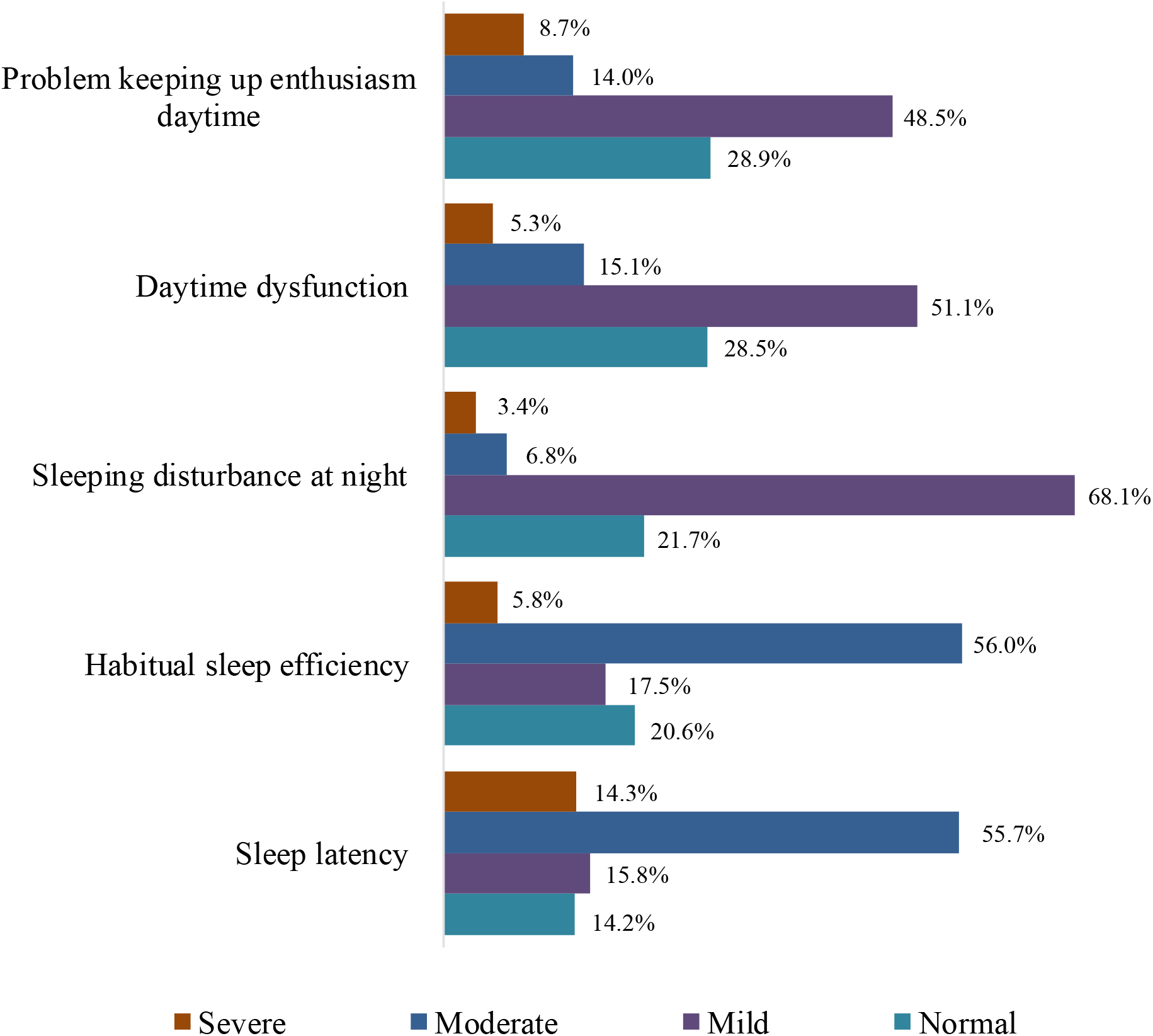
The PSQI components (n=530)

**Figure 2:**
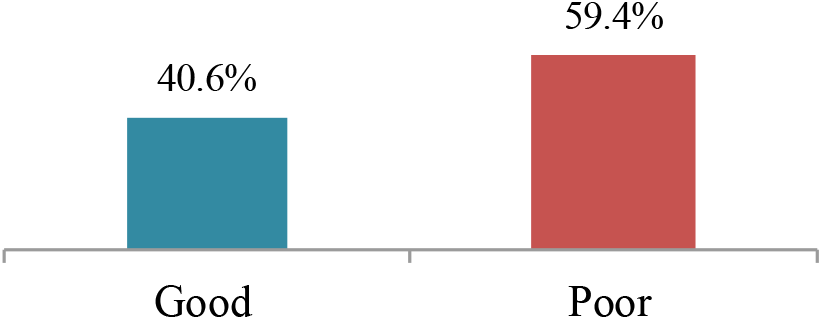
Levels of sleep quality by PSQI scale (n= 530)

**Figure 3:**
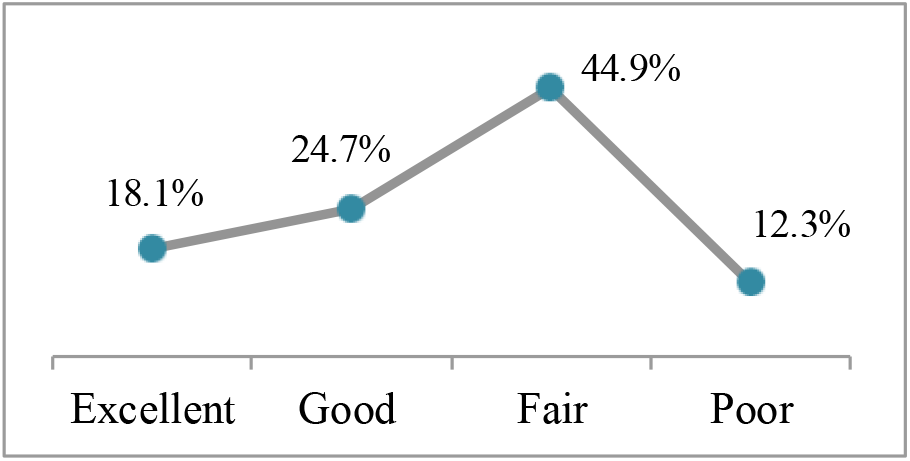
Levels of subjective sleep quality (n= 530)

A significant association was found between students’ sleep quality and socio-demographic factors such as age, family earner’s occupation, residence, family type, and household income (p<0.05). Poor sleep quality was prevalent among adolescents (82.8%), service-holder families (78.7%), rural residents (63.3%), joint families (67.1%), and households earning ≤75,000 BDT (86.7%). Significant relationships were also found between students’ sleep quality and other factors such as COVID-19 diagnosis, family member loss, and comorbidities (p<0.05). Poor sleep quality was higher among COVID-19 positive individuals (70.7%), those who lost a family member (85.0%), and those with comorbidities (75.0%).

Lifestyle factors such as class attendance, online media usage, BMI, physical exercise, tea/coffee consumption, and smoking were significantly associated with sleep quality (p<0.05). Poor sleep quality was observed in students with irregular class attendance (82.7%), reduced online media time (86.7%), obesity (90.7%), occasional exercise (76.0%), regular tea/coffee consumption (66.1%), and increased smoking (62.8%). Sleep quality factors such as daytime sleep duration, nighttime sleep latency, and sleeping pill usage in the past month were significantly associated with sleep quality (p<0.05). Poor sleep quality was observed in students who slept >2 hours during the day (64.7%), had a sleep latency of over 60 minutes at night (70.9%), and occasionally used sleeping pills (92.6%).

All components of the PSQI were significantly associated with sleep quality (p<0.05). Poor sleep quality was observed in students with severe sleep latency (85.5%), severe habitual sleep efficiency issues (100%), moderate nighttime disturbances (88.9%), severe nighttime disturbances (100%), and moderate daytime enthusiasm problems (98.4%). Subjective sleep quality was also significantly related to sleep quality (p<0.05), with poor sleep quality found in students reporting fair subjective sleep quality (97.5%).

Significant associations (p<0.05) have been found between sleep quality and mean age, monthly household income, daily study hours, BMI, daytime sleep duration, and sleep latency (Table IV).

**Table IV:**
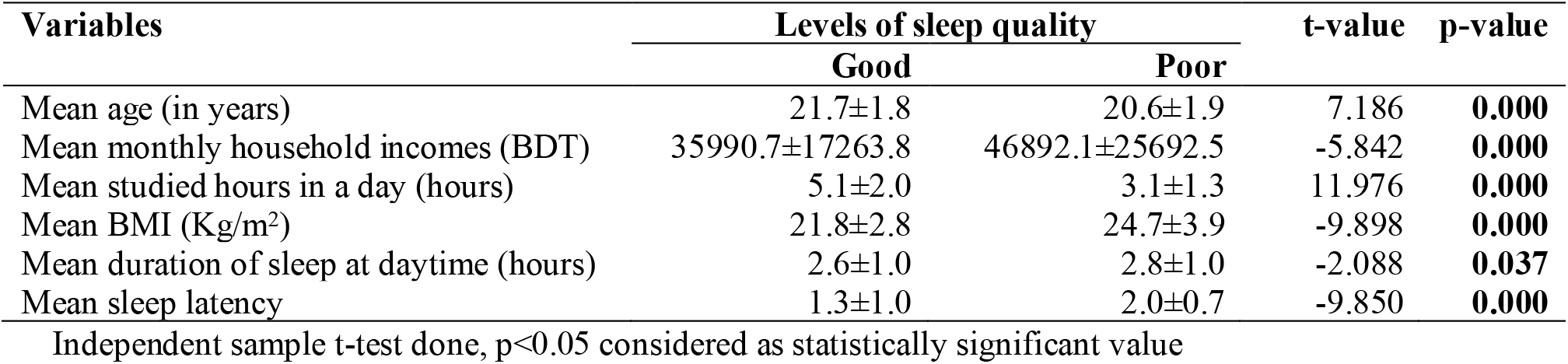
Association of factors related with the sleep quality (n=530)

The model precisely forecasts the Global PSQI score (F=17.403, p<0.001), accounting for 34.5% of the variation (R-squared= 0.366). Sleep latency, sleep quality declines with longer sleep delay (F=9.160, p<0.001). Habitual sleep efficiency, poorer sleep efficiency correlates with lower sleep quality (F=4.421, p=0.004). Sleep disturbance at night, nighttime disruptions reduce sleep quality (F= 3.657, p=0.012). Daytime dysfunction was associated with poorer sleep quality (F=6.494, p<0.001). Problem difficulty retaining excitement in the daytime predicts poorer sleep quality (F=3.690, p=0.012). Higher SSQ levels were substantially related with poor sleep quality (F=11.417, p<0.001). (Table V)

**Table V:**
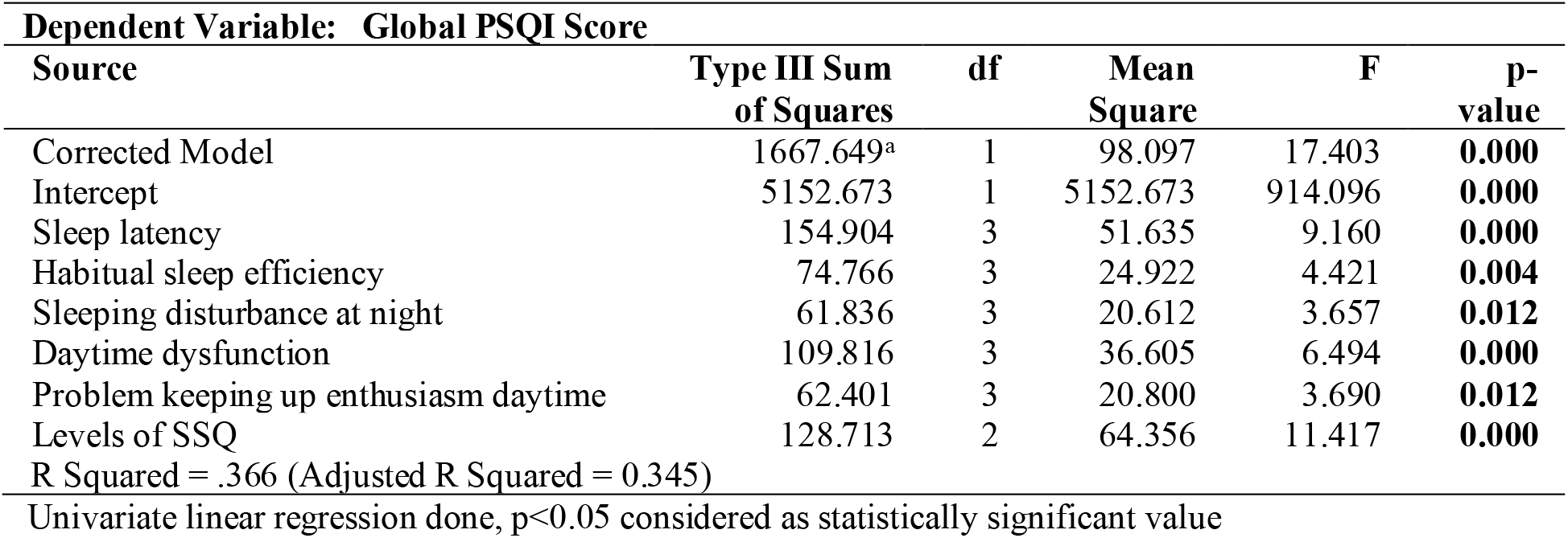
Univariate linear regression model.

| **Dependent Variable: Global PSQI Score** |  |  |  |  |  |
| --- | --- | --- | --- | --- | --- |
| **Source** | **Type III Sum of Squares** | **df** | **Mean Square** | **F** | **p-value** |
| Corrected Model | 1667.649<sup>a</sup> | 1 | 98.097 | 17.403 | **0.000** |
| Intercept | 5152.673 | 1 | 5152.673 | 914.096 | **0.000** |
| Sleep latency | 154.904 | 3 | 51.635 | 9.160 | **0.000** |
| Habitual sleep efficiency | 74.766 | 3 | 24.922 | 4.421 | **0.004** |
| Sleeping disturbance at night | 61.836 | 3 | 20.612 | 3.657 | **0.012** |
| Daytime dysfunction | 109.816 | 3 | 36.605 | 6.494 | **0.000** |
| Problem keeping up enthusiasm daytime | 62.401 | 3 | 20.800 | 3.690 | **0.012** |
| Levels of SSQ | 128.713 | 2 | 64.356 | 11.417 | **0.000** |
| R Squared = .366 (Adjusted R Squared = 0.345) |  |  |  |  |  |

Univariate linear regression done, $p < 0.05$ considered as statistically significant value

Table VI showed the logistic regression analysis revealed several significant findings regarding sleep quality. For very good sleep quality, greater sleep disturbances at night significantly increased the odds of achieving very good sleep (p=0.026). For good sleep quality, longer sleep latency (p=0.003), more sleep disturbances at night (p=0.005), greater daytime dysfunction (p=0.001), and difficulty maintaining enthusiasm (p<0.001) were all significant factors. For fair sleep quality, longer sleep latency (p<0.001), more sleep disturbances at night (p<0.001), and difficulty maintaining enthusiasm (p<0.001) were strongly associated with fair sleep quality.

**Table VI:** Logistic regression model.

| **Subjective sleep quality** |  | **B** | **Std. Error** | **Wald** | **df** | **p-value** | **Exp (B)** | **95% Confidence Interval for Exp (B)** |  |
| --- | --- | --- | --- | --- | --- | --- | --- | --- | --- |
|  |  |  |  |  |  |  |  | **Lower Bound** | **Upper Bound** |
| Excellent | Sleep latency | 0.374 | 0.406 | 0.852 | 1 | 0.356 | 1.454 | 0.657 | 3.219 |
|  | Habitual sleep efficiency | -0.631 | 0.390 | 2.619 | 1 | 0.106 | 0.532 | 0.248 | 1.143 |
|  | Sleeping disturbance at night | 0.783 | 0.352 | 4.957 | 1 | 0.026 | 2.188 | 1.098 | 4.357 |
|  | Daytime dysfunction | 0.473 | 0.309 | 2.343 | 1 | 0.126 | 1.605 | 0.876 | 2.940 |
|  | Problem keeping up enthusiasm daytime | 0.311 | 0.227 | 1.877 | 1 | 0.171 | 1.365 | 0.875 | 2.130 |
| Good | Sleep latency | 1.208 | 0.414 | 8.535 | 1 | **0.003** | 3.348 | 1.488 | 7.531 |
|  | Habitual sleep efficiency | -0.291 | 0.386 | 0.569 | 1 | 0.451 | 0.747 | 0.351 | 1.592 |
|  | Sleeping disturbance at night | 0.943 | 0.336 | 7.855 | 1 | **0.005** | 2.567 | 1.328 | 4.962 |
|  | Daytime dysfunction | 1.006 | 0.292 | 11.847 | 1 | **0.001** | 2.735 | 1.542 | 4.849 |
|  | Problem keeping up enthusiasm daytime | 0.834 | 0.221 | 14.222 | 1 | **0.000** | 2.302 | 1.493 | 3.552 |
| Fair | Sleep latency | 2.437 | 0.517 | 22.259 | 1 | **0.000** | 11.439 | 4.156 | 31.483 |
|  | Habitual sleep efficiency | -0.166 | 0.493 | 0.114 | 1 | 0.736 | 0.847 | 0.322 | 2.225 |
|  | Sleeping disturbance at night | 1.572 | 0.383 | 16.872 | 1 | **0.000** | 4.818 | 2.275 | 10.203 |
|  | Daytime dysfunction | 0.303 | 0.367 | 0.683 | 1 | 0.409 | 1.354 | 0.660 | 2.777 |
|  | Problem keeping up enthusiasm daytime | 1.494 | 0.286 | 27.226 | 1 | **0.000** | 4.453 | 2.541 | 7.803 |

The reference category is: 0 (Poor).
Logistic regression done, $p < 0.05$ considered as statistically significant value

## Discussion

This study assessed sleep quality among university students during the COVID-19 pandemic. The survey’s findings revealed a significant association between age groups and sleep quality, with the majority of students (33.6%) falling into the 22-23 age range. However, the majority of participants (64.7%) in another study conducted in Bangladesh were between the ages of 18-25 years^26^, and there was a deceptive association between adolescence and sleep quality^24,27^. Other studies found a significant relationship between sleep disorders and marital status and gender^23,28^, whereas our study found none. Our study found relationships within occupation, monthly family income, residence, and family type with sleep quality. However, relationships were observed with small household size and low socioeconomic status^2,4^. There were no discernible associates found between age, gender, marital status, and educational attainment in a survey conducted in Turkey^29^. The type of residence was also found to have an impact in other studies, participants (66.8%) felt not get enough sleep since they lived on campus^30^. Overall, 37% of Iranians living in urban areas were classified as poor sleepers^31^ and conversely, 27.7% of those in rural China reported having poor sleep quality^32^.

Significant relations were found between students’ sleep quality and factors such as being diagnosed with COVID-19 and losing family members to the virus. However, another study in Bangladesh found no correlation between sleep quality and having family members affected by COVID-19-related deaths^26^.

Comparing the PSQI score with a study conducted in Switzerland during the COVID-19 pandemic^33^, it was found that sleep quality worsened over time (p=0.035). Reports from Portugal linked home confinement without employment to sleep issues^34^, while in the United States; inadequate sleep was associated with COVID-19-related anxiety^35^. A study on university students in Lebanon showed similar results^36^, with over half of participants reporting poor sleep quality based on their PSQI scores.

Co-morbidities were significantly associated with sleep quality (p<0.05), aligning with a U.S. study linking sleep restriction to obesity and diabetes^15^. BMI was also significantly associated with sleep quality (p<0.05), with a U.S. study similarly reporting higher BMI linked to less sleep^14^. Although only 34.7% of students in this study exercised, a significant correlation with sleep quality was found (p<0.05). Conversely, a study in Lebanon found no substantial link between physical activity and PSQI, despite 56.6% of students reporting prior physical activity^36^.

Significant relations were found between students’ sleep quality, daily study hours, and online media usage (p<0.05). A study reported that emotional and intellectual stress negatively affected sleep, with 15% of students falling asleep in class weekly and 25% experiencing severe daytime sleepiness^2^. Similarly, in a survey showed that using electronic devices before sleep was significantly linked to poor sleep quality ^38^.

This survey found significant relationships between tea or coffee intake and sleep quality (p<0.05). A Saudi study reported poor sleep quality in 75.5% of daily tea drinkers and 80% of daily coffee consumers^17^, while a Lebanese study found no significant link between caffeine use and PSQI^36^. Smoking patterns increased to 67.4% during COVID-19, with a significant association between smoking and sleep quality (p<0.05). However, a Bangladeshi study found no such association^26^. In Malaysia^37^, smoking increased during COVID-19, and in other studies, nearly half of participants smoked, with a significant link between cigarette consumption and poorer sleep quality^39,40^.

This survey revealed that 73% of students slept <6 hours daily, with 44.9% reporting fair sleep quality, followed by good (24.7%), very good (18.1%), and poor (12.3%) sleep quality. Similarly, 76% of students in Saudi Arabia had poor sleep quality (mean PSQI score: 7.11±3.84)^17^. Other studies showed that 58.7% of students reported poor sleep quality^8^, a trend also noted in the USA^2^. In Taiwan, the average daily sleep time was 6.7±1.3 hours^16^, while students in Lebanon had an average PSQI score of 6.57±3.49.^36^

## Conclusion

In conclusion, the study identified key factors influencing sleep quality among university students. Poor sleep quality was associated with demographic factors like age, family income, and residence, as well as health issues such as COVID-19, family loss, and comorbidities. Lifestyle behaviors, including irregular class attendance, high online media use, obesity, lack of exercise, and smoking, were also linked to poor sleep. Furthermore, longer sleep latency, daytime sleep, and sleeping pill usage contributed to lower sleep quality. It also underlined that sleep disturbances, daytime dysfunction, and difficulty maintaining enthusiasm were significant predictors of sleep quality. Addressing these factors could help improve sleep quality in this population.

## Data Availability

All data produced in the present study are available upon reasonable request to the authors

## Author’s contributions

Conceptualization, methods and literature reviews: Nurunnabi M and Abbas MG; Data collection: Abbas MG; Statistical analysis: Nurunnabi M; Preparation of draft manuscript: Nurunnabi M, Khan MK, Tabassum TT, Farha T, and Akter F; and Finalization of manuscript: Nurunnabi M, Khan MK, Kaiser FR, Abbas MG, Tabassum TT, Farha T, Akter F, and Tarafdar MA. All the authors approved the final manuscript

## Artificial Intelligence Declaration

AI (Chat-GPT 4) was used to enhance the clarity of the manuscript and assist with language editing

## Acknowledgments

The authors gratefully acknowledge the students and data collectors for their valuable contributions to this study.

## References

1. Lund HG, Reider BD, Whiting AB, Prichard JR. Sleep patterns and predictors of disturbed sleep in a large population of college students. Journal of Adolescent Health. 2010;46(2):124–32.

2. Dregan A, Armstrong D. Adolescence sleep disturbances as predictors of adulthood sleep disturbances-A cohort study. Journal of Adolescent Health. 2010;46(5):482–7.

3. Ahmed F, Ara J, Nurunnabi M, Akter F. Insomnia and Associated Social Factors among Medical Support Staff during the Covid-19 Pandemic. Mymensingh Medical Journal. 2025; 34(1):234–242.

4. Mostarin S, Haque A, Alam MR, Choudhury R, Nurunnabi M, Sultana H, Abbas MG. Sleep Pattern of Undergraduate Medical Students of the selected Medical Colleges in Dhaka during COVID-19 Pandemic: An Online Survey. Z H Shikder Women’s Medical College Journal. 2022;4(2):4 –10.

5. Buysse DJ, Reynolds III CF, Monk TH, Berman SR, Kupfer DJ. The Pittsburgh Sleep Quality Index: a new instrument for psychiatric practice and research. Psychiatry Research. 1989;28(2):193–213.

6. Mollayeva T, Thurairajah P, Burton K, Mollayeva S, Shapiro CM, Colantonio A. The Pittsburgh sleep quality index as a screening tool for sleep dysfunction in clinical and non-clinical samples: A systematic review and meta-analysis. Sleep Medicine Reviews. 2016;25:52–73.

7. Abbas MG, Mostarin S, Rimmi SH, Sultana R, Hossain S, Nurunnabi M. Sleep Quality among Undergraduate Public Medical and University Students: A Comparative Study. KYAMC Journal. 2023;13(4):223–228.

8. Peltzer K, Pengpid S. Sleep duration and health correlates among university students in 26 countries. Psychology, Health & Medicine. 2016;21(2):208–20.

9. Schlarb AA, Kulessa D, Gulewitsch MD. Sleep characteristics, sleep problems, and associations of self-efficacy among German university students. Nature and Science of Sleep. 2012:1–7.

10. Prather AA, Janicki-Deverts D, Hall MH, Cohen S. Behaviorally assessed sleep and susceptibility to the common cold. Sleep. 2015;38(9):1353–9.

11. Goel N, Rao H, Durmer JS, Dinges DF. Neurocognitive consequences of sleep deprivation. In Seminars in Neurology 2009;29(4):320–39.

12. Walker MP. The role of sleep-in cognition and emotion. Annals of the New York Academy of Sciences. 2009;1156(1):168–97.

13. Baglioni C, Battagliese G, Feige B, Spiegelhalder K, Nissen C, Voderholzer U, Lombardo C, Riemann D. Insomnia as a predictor of depression: a meta-analytic evaluation of longitudinal epidemiological studies. Journal of Affective Disorders. 2011;135(1-3):10–9.

14. Taheri S, Lin L, Austin D, Young T, Mignot E. Short sleep duration is associated with reduced leptin, elevated ghrelin, and increased body mass index. PLoS Medicine. 2004;1(3):e62.

15. Buxton OM, Cain SW, O’Connor SP, Porter JH, Duffy JF, Wang W, Czeisler CA, Shea SA. Adverse metabolic consequences in humans of prolonged sleep restriction combined with circadian disruption. Science Translational Medicine. 2012;4(129):129–43.

16. Kang JH, Chen SC. Effects of an irregular bedtime schedule on sleep quality, daytime sleepiness, and fatigue among university students in Taiwan. BMC Public Health. 2009;9:1–6.

17. Almojali AI, Almalki SA, Alothman AS, Masuadi EM, Alaqeel MK. The prevalence and association of stress with sleep quality among medical students. Journal of Epidemiology and Global Health. 2017;7(3):169–74.

18. Kessler RC, Foster CL, Saunders WB, Stang PE. Social consequences of psychiatric disorders, I: Educational attainment. American Journal of Psychiatry. 1995;152(7):1026–32.

19. Nurunnabi M, Tarafdar MA, Begum A, Jahan S, Islam AFMR. Adolescent Suicide and Suicidal Behavior: A Review. Z H Shikder Women’s Medical College Journal. 2021;3(2):38–42.

20. Khan F, Haroon H, Murtaza H, Anwar E. Determinants of sleep quality among undergraduate students of universities of Karachi. Annals of Psychophysiology. 2016:4–13.

21. Rahman MR, Akhtar K, Azizi S, Nurunnabi M, Chowdhury MAM, Adnan MA. Quality of Life of COVID-19 Patients Attending in Selected Post-COVID Units. Journal of Preventive and Social Medicine. 2023;42(1):49–53.

22. Nurunnabi M, Tarafdar MA, Islam MS, Uddin NMM, Akter F, Begum A. Demographic Trends of Adolescent Suicidal Hanging Deaths in Sylhet. Z H Shikder Women’s Medical College Journal. 2024;6(1):21–25.

23. Nurunnabi M, Nazia A, Chowdhury N, Alam MB, Islam MM, Kakoly NS. Prevalence of Post-traumatic stress disorder among Physicians during the COVID-19 Pandemic. Bangladesh Medical Journal. 2022;51(1):52–58.

24. Ahmed Z, Ahmed MM, Ali MA, Nurunnabi M, Hossain S, Abbas MG, Lima FA, Hossain TB. Impact of Coronavirus Infection on Adolescence during the COVID-19 Pandemic. IAHS Medical Journal. 2022;5(1):3–7.

25. Islam S, Akter R, Sikder T, Griffiths MD. Prevalence and factors associated with depression and anxiety among first-year university students in Bangladesh: a cross-sectional study. International Journal of Mental Health and Addiction. 2020:1–4.

26. Islam MS, Rahman ME, Zubayer AA, Bhuiyan MR, Khan MK, Hossain L, Sujon MM. Investigating poor sleep quality and associated factors during the COVID-19 pandemic: A population-based survey in Bangladesh. Frontiers in Public Health. 2021;9:724520.

27. Nurunnabi M, Tarafdar MA, Islam MS, Uddin NMM, Akter F, Begum A. Demographic Trends of Adolescent Suicidal Hanging Deaths in Sylhet. Z H Shikder Women’s Medical College Journal. 2024;6(1):21–25.

28. Jahan SM, Hossain SR, Sayeed UB, Wahab A, Rahman T, Hossain A. Association between internet addiction and sleep quality among students: a cross-sectional study in Bangladesh. Sleep and Biological Rhythms. 2019;17:323–9.

29. Akinci T, BaŞar HM. Relationship between sleep quality and the psychological status of patients hospitalized with COVID-19. Sleep medicine. 2021;80:167–70.

30. Qin P, Brown CA. Sleep practices of university students living in residence. International Journal of higher education. 2017;6(5):14–25.

31. Asghari A, Kamrava SK, Ghalehbaghi B, Nojomi M. Subjective sleep quality in urban population. Archives of Iranian medicine. 2012;15(2):95–8.

32. Dong X, Wang Y, Chen Y, Wang X, Zhu J, Wang N, Jiang Q, Fu C. Poor sleep quality and influencing factors among rural adults in Deqing, China. Sleep and Breathing. 2018;22:1213–20.

33. Targa AD, Benítez ID, Moncusi-Moix A, Arguimbau M, de Batlle J, Dalmases M, Barbe F. Decrease in sleep quality during COVID-19 outbreak. Sleep and Breathing. 2021;25:1055–61.

34. Pinto J, van Zeller M, Amorim P, Pimentel A, Dantas P, Eusébio E, Neves A, Pipa J, Santa Clara E, Santiago T, Viana P. Sleep quality in times of Covid-19 pandemic. Sleep medicine. 2020;74:81–5.

35. Hyun S, Hahm HC, Wong GT, Zhang E, Liu CH. Psychological correlates of poor sleep quality among US young adults during the COVID-19 pandemic. Sleep medicine. 2021;78:51–6.

36. Kabrita CS, Hajjar-Muca TA, Duffy JF. Predictors of poor sleep quality among Lebanese university students: association between evening typology, lifestyle behaviors, and sleep habits. Nature and Science of Sleep. 2014:11–8.

37. Aye LM, Lee WH. Poor sleep quality and its associated factors among working adults during COVID-19 pandemic in Malaysia. Cambridge Prisms: Global Mental Health. 2024;11:e28.

38. Garett R, Liu S, Young SD. The relationship between social media use and sleep quality among undergraduate students. Information, Communication & Society. 2018;21(2):163–73.

39. Vail-Smith K, Felts WM, Becker C. Relationship between sleep quality and health risk behaviors in undergraduate college students. College student journal. 2009;43(3):924–30.

40. Seun-Fadipe CT, Mosaku KS. Sleep quality and psychological distress among undergraduate students of a Nigerian university. Sleep Health. 2017;3(3):190–4.

